# Minimum manufacturing costs, national prices and estimated global availability of new repurposed therapies for COVID-19

**DOI:** 10.1101/2021.06.01.21258147

**Authors:** Junzheng Wang, Jacob Levi, Leah Ellis, Andrew Hill

## Abstract

**Background:** Currently, only dexamethasone, tocilizumab and sarilumab have conclusively been shown to reduce mortality of COVID-19. Safe and effective treatments will need to be both affordable and widely available globally to be used alongside vaccination programmes. This analysis will estimate and compare potential generic minimum costs of a selection of approved COVID-19 drug candidates with available international list prices.

**Methods:** We searched for repurposed drugs that have been approved by at least one of the WHO, FDA or NICE, or at least given emergency use authorisation or recommended for off-label prescription. Drug prices were searched for, for dexamethasone, budesonide, baricitinib, tocilizumab, casirivimab and imdevimab, and sarilumab using active pharmaceutical ingredients (API) data extracted from global shipping records. This was compared with national pricing data from a range of low, medium, and high-income countries. Annual API export volumes from India were used to estimate the current availability of each drug.

**Results:** Repurposed therapies can be generically manufactured for some treatments at very low per-course costs, ranging from $2.58 for IV dexamethasone (or $0.19 orally) and $4.34 for inhaled budesonide. No export price data was available for baricitinib, tocilizumab, casirivimab and imdevimab or sarilumab, but courses of these treatments are priced highly, ranging from $6.67 for baricitinib to $875.5 for sarilumab. When comparing international list prices, we found wide variations between countries.

**Conclusions:** Successful management of COVID-19 will require equitable access to treatment for all populations, not just those able to pay high prices. Dexamethasone and budesonide are widely available and affordable, whilst monoclonal antibodies and IV treatment courses are more expensive.

**Key Points:**

- Re-purposed drugs must be affordable worldwide to compliment COVID-19 vaccine programmes.
- Estimated costs/course were: dexamethasone (Oral $0.22, IV $2.58), budesonide ($4.34), baricitnib ($6.67), tocilizumab ($410.59), sarilumab ($875.70). Casirivimab and imdevimab = no data available.
- High drug prices will limit access.

## Introduction

18-months from the initial COVID-19 outbreak, drug repurposing with intention to treat and prevent infections is still an important strategy alongside vaccine rollout. The uneven global vaccine deployment, “vaccine nationalism” [1], difficulties caused by vaccine hesitancy and emerging new variants of concern all means that an excessively vaccine-dependent strategy may not be fully effective. Even in countries with high vaccine coverage, there are still symptomatic patients who may benefit from treatment options. For many developing countries, mass vaccination required to reduce the significant healthcare burden of COVID-19 may remain unattainable for several years.

Treatments, when they emerge from the vast number of clinical trials currently underway, will need to overcome the same barriers of efficacy, safety, affordability, and availability as vaccines. In our 2020 analysis of potential costs of generic production for drug candidates then under active investigation [2], we proposed four key tests:

1. *Efficacy –* drugs should demonstrate therapeutic clinical efficacy and/or prophylactic properties.
2. *Safety* – drugs should be safe and well tolerated in all populations with minimal side effects or drug interactions.
3. *Affordability* – drugs should be widely accessible as low-cost generics so that all populations can equitably afford the cost of treatment.
4. *Availability –* drugs must be readily available or have the potential for rapid up-scaled production to meet global demand.

Some treatments have passed the first and second hurdle; dexamethasone, remdesivir, tocilizumab and sarilumab are WHO approved, whilst the USA FDA has also granted an emergency use authorisation for baricitinib and the UK NICE has given advise for off-label prescriptions of inhaled budesonide in certain patients with COVID-19. Many previously investigated drugs however have now been proven ineffective. Large-scale studies such as SOLIDARITY and RECOVERY have found previously leading candidates such as hydroxychloroquine [3], lopinavir/ritonavir [4,5], subcutaneous interferon beta-1a [4] and azithromycin [6] to have no clinical benefit. Additionally, no evidence of efficacy was found for colchicine in the RECOVERY trial [7] despite some preliminary evidence of efficacy from the published COLCORONA trial [8]. Furthermore, despite studies with fraudulent data attempting to portray ivermectin as a promising treatment, meta-analysis has demonstrated no clinical benefit for ivermectin [9,10]. Despite FDA approval, remdesivir use has led to shorter hospitalisation times, but with no significant mortality benefits [4].

Baricitinib, a janus kinase (JAK) inhibitor being tested in 14 studies amongst 56,168 patients and is mostly being given together with remdesivir [11]. It has been granted an emergency use authorisation by the FDA [12]. More recently, promising press reports have claimed a 50% reduction in the risk of hospitalisation or death (from 14% to 7%) for the re-purposed anti-viral therapy molnupiravir, but full peer reviewed data is not yet available [13] and the FDA, NICE nor WHO have given approval yet (as of October 10^th^, 2021). Furthermore, the antibody cocktail casirivimab and imdevimab has been shown to reduce COVID-19 related hospitalisation or all cause death in small-randomised studies for seronegative patients [14,15] and has received an EUA from the FDA and recent approval by NICE for seronegative patients [16].

However, questions of affordability and availability should be equally important as efficacy and safety. Treatments are of limited value if their supply is limited and the cost prohibitive, as in the case of tocilizumab and sarilumab. These monoclonal antibody interleukin-6 (IL-6) receptor inhibitors were found to improve outcomes in patients with severe COVID-19 [17], but cost several hundred dollars per dose, and require complex bioreactors to produce. Dexamethasone, the first treatment found to increase survival and is widely available as a low-cost generic, is only applicable for patients requiring oxygen supplementation [18]. An effective anti-viral agent for asymptomatic, mild, or moderate patients that limits progression and treats COVID-19 is yet to be approved.

We analysed their costs of production based on cost of active pharmaceutical ingredients (API) for drugs that have been approved by regulatory bodies, and offer a snapshot of global API availability, and current list prices in a range of countries where data was available. Given the limited transparency on drug costs and prices, this study aimed to contribute to the current evidence base to support public health decision makers should any analysed drug be proven effective against COVID-19.

## Methods

### Drug selection

Clinical trial registries such as clinicaltrials.gov and covid-nma.com were screened alongside current published literature to identify promising repurposed drug candidates. Only those approved by the USA FDA, the WHO, or the UK NICE or at least given emergency use authorisation or recommended for off label use were included in the final analysis. The Panjiva database [19] and national drug price sources were searched for available export data (Appendix 1). Remdesivir was not re-analysed in this paper as it was assessed in our previous publication [2].

### Oral medications

Methodologies for estimating minimum costs of oral medication production were previously published [2]. Costs of Indian API exports were analysed (JW and JL) and independently reviewed (LE) using the online tracking database Panjiva, which showed API shipment details including quantities and costs per kilogram. All available API data from 2016 to June 2021 were included. For all oral drugs we excluded shipments <1kg in volume and excluded the 15% highest and lowest priced shipments were removed to exclude outliers. For oral drugs, 5% API loss during tableting process, and a conversion cost of $0.01 per tablet was factored into calculations, alongside a multiplier based on API mass to account for the expense of excipients (additional substances needed to convert API into the finished pharmaceutical product). Profit margins of 10% and Indian taxation of 27% on profit were added. Indian production was assumed given the country’s leading role in global generic medicine manufacturing [20].

### IV dexamethasone

For injectables, an adjusted published methodology was followed to account for the cost of ampules, additional transport costs, and projected 20% API loss during formulation and filling [2,21].

### Inhaled budesonide

In the STOIC and PRINCIPLE studies, patients were asked to use two puffs of a 400mcg inhaler twice daily, (800mcg BD) for 14 or 28 days, with median duration being seven days [22, 23]. Cost per kilogram of budesonide dry powder API was estimated using the same methodology above, before accounting for the cost of inhaler devices (Pulmicort turbohaler $3), additional transport costs, and a projected 20% API loss during formulation and filling. The following alternative inhalers are also permitted for use by NICE; Pulmicort Turbohaler 200 micrograms, Budelin Novolizer 200 micrograms, Easyhaler Budesonide 400 micrograms and Easyhaler Budesonide 200 micrograms.

### Tocilizumab, sarilumab, baricitinib

No data for the cost or volume of monoclonal antibodies production were available, as this depends heavily on the efficiency and size of bioreactors used [24]. Also, no API data was available for baricitinib or casirivimab and imdevimab. Therefore, list prices in a range of countries, in particular developing economies, were tracked as a proxy, as it is assumed that the generic medicine could be reasonably produced at this level or less. Unfortunately, no national price data was yet available for casirivimab and imdevimab. There are news reports of tocilizumab and sarilumab being sold in Indian black markets at around $658.50 per 600mg dose [25].

### International list price comparison

Estimated minimum production costs were compared with published list prices in a range of countries across the economic development spectrum to give a representative sample of prices globally. Data sources by country are detailed in Appendix 2. Data was collected and reviewed independently by JL and LE, intended to offer a snapshot at time of writing.

For consistency, a single data source per country was used for all searches of drug prices, based on the organisation of data and perceived reliability except for the USA. For the United States, we used the commercial pharmacy (USA, Pharm) cost as well as public costs from the veterans’ affairs system (USA, Vets) listed separately. This is because the USA has a healthcare system with a wide range of insurance options, and the USA veterans’ affairs programme insures many millions of Americans, whilst millions more Americans are uninsured and must pay out of pocket for most medications. Not all drugs analysed were available in every country, and in some countries without official databases, online pharmacy sites were used instead. Where several prices were available in the same database, the lowest reliable price was selected.

### API availability

Panjiva data was used to analyse API export volumes per drug from India and estimate the maximum number of treatment courses potentially producible over a 12-month period.

### Key assumptions

For some drugs, such as dexamethasone and tocilizumab, which are dosed on a per kilogram basis we used an average adult body mass of 70kg.

The costs of regulatory filings and approvals are often significant add-ons to the initial use of drugs in any specific country. All drugs analysed in this study have been approved for treatment for some indications already. We have not included the cost or timing associated with regulatory approvals for the use of these drugs. We assume that the WHO and other influential regulatory agencies will cooperate to define a pathway for use of these drugs which does not include additional financial outlays or filing for marketing approvals.

## Results

The results are summarised in Table 1.

**Table 1.**
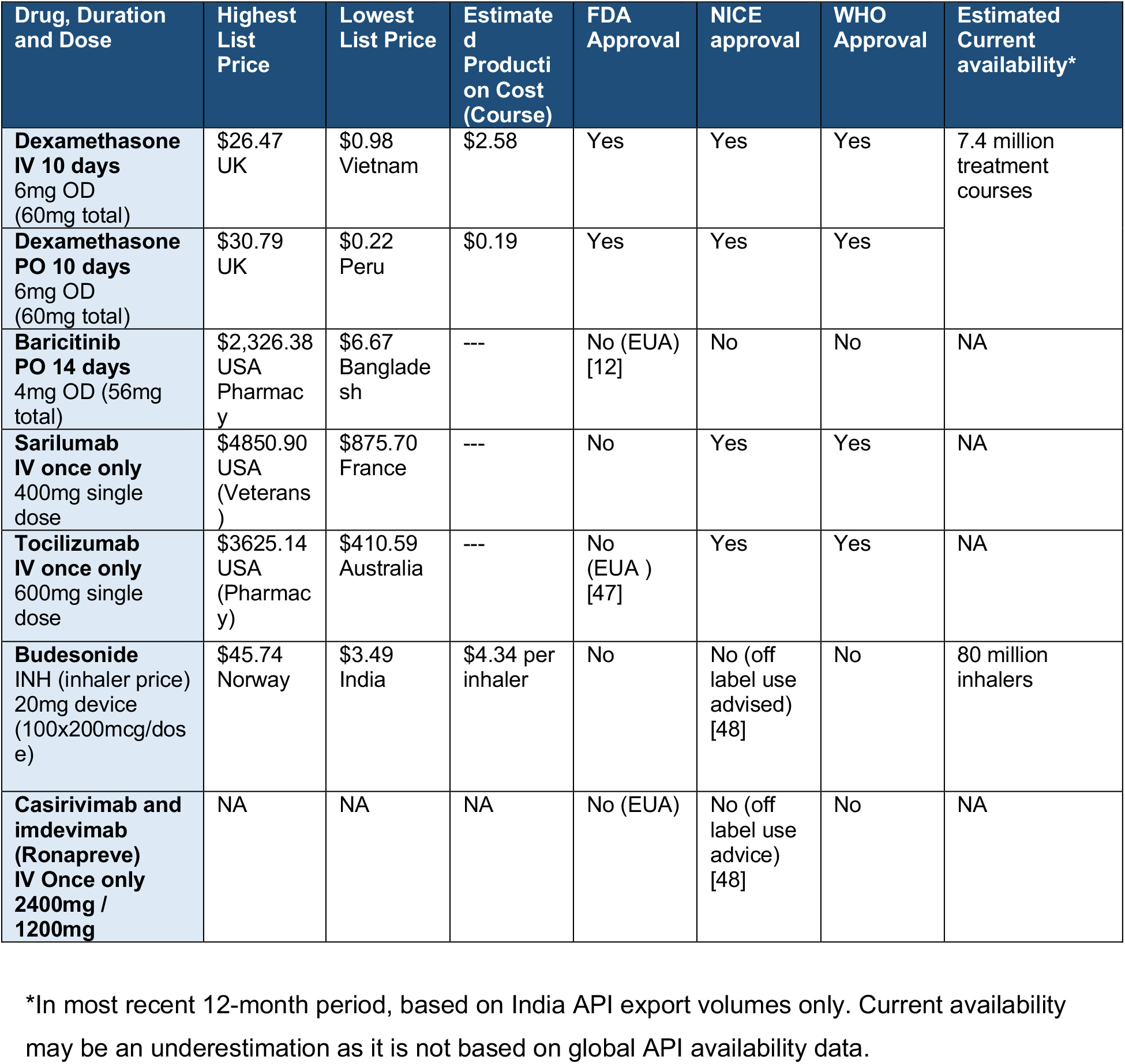
Summary of repurposed drug prices for COVID-19 and estimated availability. Summary of list prices, estimated production costs, and current availability of potential COVID-19 drugs selected for analysis. OD = Once daily, BD = twice per day, EUA = Emergency Use Authorisation, (baricitinib only has EUA to be given in combination with remdesivir)

| Drug, Duration and Dose | Highest List Price | Lowest List Price | Estimated Production Cost (Course) | FDA Approval | NICE approval | WHO Approval | Estimated Current availability* |
| --- | --- | --- | --- | --- | --- | --- | --- |
| **Dexamethasone IV 10 days**<br>6mg OD (60mg total) | \$26.47 UK | \$0.98 Vietnam | \$2.58 | Yes | Yes | Yes | 7.4 million treatment courses |
| **Dexamethasone PO 10 days**<br>6mg OD (60mg total) | \$30.79 UK | \$0.22 Peru | \$0.19 | Yes | Yes | Yes | |
| **Baricitinib PO 14 days**<br>4mg OD (56mg total) | \$2,326.38 USA Pharmacy | \$6.67 Bangladesh | --- | No (EUA) [12] | No | No | NA |
| **Sarilumab IV once only**<br>400mg single dose | \$4850.90 USA (Veterans) | \$875.70 France | --- | No | Yes | Yes | NA |
| **Tocilizumab IV once only**<br>600mg single dose | \$3625.14 USA (Pharmacy) | \$410.59 Australia | --- | No (EUA) [47] | Yes | Yes | NA |
| **Budesonide INH (inhaler price)**<br>20mg device (100x200mcg/dose) | \$45.74 Norway | \$3.49 India | \$4.34 per inhaler | No | No (off label use advised) [48] | No | 80 million inhalers |
| **Casirivimab and imdevimab (Ronapreve) IV Once only**<br>2400mg / 1200mg | NA | NA | NA | No (EUA) | No (off label use advice) [48] | No | NA |

\*In most recent 12-month period, based on India API export volumes only. Current availability may be an underestimation as it is not based on global API availability data.

### Dexamethasone

The widely used corticosteroid, dexamethasone has an economy of scale which supports low-cost production. Current national guidelines for dexamethasone treatment in COVID-19 most commonly recommend a 7-10-day courses of 6mg oral or IV [26, 27]. Shorter courses are used if patients improve or are discharged early, and sometimes longer courses and higher doses are used in patients with ongoing fibrosis and hyper-inflammation confirmed by high-resolution CT imaging.

This analysis used a 10-day, 6mg/day course. The weighted-mean API cost between 2016-2020 was estimated to be $1116/kg; after factoring in the costs of excipients, formulation, tax, and profit, a 10-day IV course could be reasonably producible for around $2.58 and around $0.19 as tablets (Figs.1a,1b). The higher cost of injectables reflect the potential for greater loss during production, cost of vials and increased cost of transport. Furthermore, significant non-drug costs associated with IV administration, including staff time, expendables required such as sterile water/syringes for reconstitution, and saline/IV lines for infusion were not included.

Comparatively, current list prices for oral dexamethasone were found to range from around $30.79 and $20.69 per course in UK and Israel respectively, to only $0.26 and $0.22 in China and Peru. For IV dexamethasone, course costs ranged between $26.47 (UK) and $0.98 (Vietnam) (Figs.2a, 2b). Sufficient API export shipments were identified in the most recent 12-month period to form 7.4 million courses, a substantial decrease on the four-year average between 2016-2019 at a theoretical 34.5 million 10-day courses per annum.

### Tocilizumab and sarilumab

Dosages for both monoclonal antibodies are based on current UK guidelines [28]. Both are given as single dose IV infusions: sarilumab at 400mg, and tocilizumab at 8mg/kg (800mg maximum). In rare circumstances, a second tocilizumab dose can be given 12-24 hours later, depending on response to first dose. This analysis is based a single infusion of sarilumab 400mg, and tocilizumab 600mg which are guideline dose-banding suggestions for bodyweights 66kg-80kg inclusive.

International list prices per single dose of sarilumab ranged between $4850.90 (USA) to $875.70 (France); whilst for tocilizumab, single dose costs ranged between $3,625.14 (USA) to $410.59 (Australia) (Figs.2c, 2d).

### Baricitinib

Baricitinib is an anti-rheumatic drug, which in clinical trials for COVID-19 [11] is dosed at 4mg/day for 14 days orally. No API data is currently available. International list prices for a 14-day treatment course were found to range between $2,326 (USA) to $6.67 (Bangladesh) (Fig.2e).

### Budesonide

The STOIC study used a protocol of 800mcg BD up to 28 days, with a median duration of seven days [22,23]. Clinically, patients generally receive individual inhaler devices containing 60-200 doses of 400mcg/dose, 200mcg/dose, or 100mcg/dose which is more than sufficient for the proposed treatment duration.

Therefore, production cost of dry power budesonide inhalers containing 20mg of API was estimated to be $4.34 (100 doses of 200mcg puffs) or $4.48 if containing 40mg of API (100 doses of 400mcg puffs), based on a weighted-mean API cost of $4325/kg. This was after accounting for the cost of the inhaler device ($3), a 20% potential API loss during production, transport costs, profit, and tax (Fig.1c). International list prices vary by country depending on the availability of the 200mcg/dose or 400mcg/dose inhaler devices and are shown in Fig.2f.

API export from India in the most recent 12-month period was estimated to be sufficient for 80 million inhalers, whilst the five-year average was estimated to be around 88 million inhaler equivalents, should the entire API production volume be used for inhalers.

## Discussion

Repurposed drugs with available API data could all be produced generically at very low cost. Oral tablets such as dexamethasone, and baricitinib can potentially all be sold for $7 to less than $1 per course, whilst inhaled budesonide could cost <$4. Intravenous drugs were more expensive, ranging from $1/course of IV dexamethasone to $411 for tocilizumab and $875 for sarilumab. Indeed, list prices in developing economies such as Bangladesh, Kenya and India are already sometimes less than our estimated costs. API costs were previously found to be a key determinant of generic medicine prices - in a 2015 study of sofosbuvir/daclatasvir, an antiviral combination for hepatitis-C, API costs accounted for 65-95% of final drug prices in competitive markets [29]. No data was yet available for casirivimab and imdevimab but we anticipate this neutralising monoclonal antibody cocktail is likely to be highly priced as and when it becomes available.

Analysis of list prices, although only a snapshot, demonstrated large variations between countries, especially for monoclonal antibodies and highlights again the importance of international global health coordination to ensure accessibility. Public-private partnerships such as the Global Fund showed that it is possible to offer drug access at near cost-price through voluntary mechanisms. Strategies such as pooled procurement, differential pricing and voluntary licenses should be utilised more widely, as over time these have become more exclusionary and available only to a shrinking list of low- and middle-income countries (LMICs). Studies of differential or tiered pricing have shown that they can result in higher prices for MICs due to arbitrary market segmentation [30].

Other tools pertinent to expanding access to medicines in the current pandemic includes the use of TRIPS flexibilities by national governments and civil society, particularly of compulsory licensing and patent oppositions. On multiple occasions this proved to be effective in addressing high prices on medicines protected by intellectual property (IP) monopolies and triggering radical price reductions [31]. Given the pandemic situation, the so-called TRIPS waiver proposal at the WTO would be a crucial initiative for removing IP barriers to access generic and/or biosimilar medicines based on cost of manufacturing [32].

In terms of availability, dexamethasone, ivermectin, budesonide and fluvoxamine and are already available as generic medicines with a large volume of API being produced already. If promising results are demonstrated for any of these drugs in large studies, they could become an option earlier on in the disease process and production must be rapidly upscaled to meet demand. Regulators and decision makers should plan for this possibility and achieve accelerated deployment especially given that the safety profiles of these drugs are already established. An important aspect will be to protect supplies for existing patients treated for indicated conditions, for example parasitic infections and rheumatoid arthritis.

Neither steroids, monoclonal antibodies, nor IL-6 receptor antagonists will be a silver bullet against COVID-19. Dexamethasone has shown a small improvement in survival for treatment of oxygen-dependent, hospitalised patients (18%), with a larger benefit seen for the treatment of mechanically ventilated patients (36%) [18]. A recent meta-analysis found a small, improved survival benefit for tocilizumab in the treatment of hospitalised patients in high income countries [33]. Conclusive benefits for other outcomes were inconclusive. It is unclear whether the same survival benefit would be seen in LMIC settings where infection control can be more challenging in immunocompromised patients. It is also unclear whether sufficient supplies of tocilizumab or sarilumab could be manufactured to meet worldwide demand.

A key barrier to regulatory decision-making on repurposed drugs has been concerns regarding quality of evidence, highlighted by the latest WHO guidelines on ivermectin [34]. A recent meta-analysis of 24 ivermectin trials included far fewer in the main analysis due to high risks of bias [9]. Funders and regulators should take a greater co-productive role with researchers to ensure clinical trials are well designed, adequately powered, and acceptable to the regulatory approval process when they are completed.

Overpriced treatments may harm already stretched healthcare systems and as is often the case with healthcare and epidemiology prevention is generally more cost effective that treatment. Vaccines remain very effective and relatively cheap per person. Despite this, the USA has spent over $1billion pre-ordering around 1.7 million courses of molnupiravir, costing an estimated $1200 per course [35]. Furthermore, the price of a 5-day remdesivir course in the USA is approximately $2,335.50 despite offering no mortality reduction and not currently being recommended by WHO.

### Limitations

This study presumed Indian generic production [20] with lower associated costs and no capital investment in new, dedicated facilities; however, following supply chain disruptions as seen earlier on in the pandemic, and currently with vaccines, countries may accept additional upfront costs and overheads for greater supply security in establishing domestic production. Fluctuating exchange rates and confidential in-country negotiated discounts for public healthcare providers means the prices we report are subject to change and are a snapshot of publicly available information at the time of writing [36]. The re-purposed drugs landscape is also a rapidly evolving pipeline, and many of the drugs we initially analysed were subsequently shown to be ineffective. Drug prices can evolve quite erratically as supply and demand, media hype and drug approval processes exert significant pressure on pharmaceutical markets and supply chains.

API production costs are subject to changes in supply and demand and optimisation of manufacturing processes. Further, analysis was dependent on Indian custom records from Panjiva. Data in other countries were unavailable so the real volume of global API production may be higher. Finally, regulatory approval costs were not considered, assuming that regulatory agencies such as WHO and FDA would coordinate rapid, emergency deployment of drugs without significant burden [37-39].

Our estimates do not include research and development costs. However, most of the investment for drug repurposing in COVID-19 has originated from public or philanthropic sources, with significant amounts of research funded directly by governments, universities, public institutions, and non-governmental organisations. A recent estimate suggested that governments have spent almost $100 billion in the last year solely on vaccine research [40].

### Aspects for further research

As more information becomes available, it is possible that other promising drugs such as imatinib, infliximab or molnupiravir may be approved by the WHO if initial clinical results from pre-prints are confirmed in larger peer reviewed RCTs. A recently published study (October 2021) has found that the 5-day course of the anti-viral molnupirivir taken at 800mg BD could profitably be sold for less than $20 per person when calculated using Panjiva API export data [41] and using raw ingredients and a synthetic process with solvent capture this could drop to below $5 per course [42]. When we ran a preliminary analysis of API Panjiva data of molnupiravir in June 2021, we found a treatment course would cost more than x10 fold higher. This illustrates just how quickly small molecule nucleoside analogues can drop in price when production is upscaled. It is imperative that new and repurposed drugs are made available at fair, near-cost of production prices, to improve access and availability for all. When data is available for casirivimab and imdevimab, we would plan to analyse their prices.

There are good global accessibility and availability for steroids and budesonide. High prices of JAK inhibitors and monoclonal antibodies are barriers for patient access in healthcare systems requiring out-of-pocket co-payments or LMIC settings. The entry of biosimilars can lead to significant price reductions, even if not on the scale of chemical medications [43,44]. One reason for higher biosimilar costs could be the overly cumbersome regulatory pathway [45] that many have argued for reform. If greater emphasis is placed on encouraging biosimilar entry, prices for tocilizumab and sarilumab could be much lower and allow for greater availability.

Historically, combination therapy proved most effective against HIV/AIDS, hepatitis-C and TB - a similar approach might be needed for COVID-19 [46]. Combinations such as antivirals given concurrently with anti-inflammatory drugs or at different disease stages may prove more effective than monotherapy and warrants further investigation. Occasionally, generic combination therapies can cost less than the sum of their parts due to increased efficiency in formulation, tableting and packaging.

## Conclusion

Currently, more conclusive data on clinical efficacy is required for most repurposed drugs being trialled for COVID-19. This analysis shows that once efficacy is demonstrated, oral repurposed drugs can be manufactured at very low costs, whilst IV treatments are a little more costly, the monoclonal anti-body treatments are expensive. In the short term until vaccination access is more widely available and uptake has improved, repurposed drugs continue to potentially offer affordable and accessible treatment options at all stages of disease, from pre-exposure prophylaxis to treatment of asymptomatic and mild infections, through to critical care.

## Data Availability

Data used for this study is available upon request to the authors.

## Acknowledgements

The authors would like to thank members of the International COVID-19 Research Group, Kajal Bhardwaj and Sergey Kondratyuk for their support of this study and their contributions towards the development of this manuscript including insights, feedback, and proof-reading.

The authors would also like to express their gratitude to Professor Joe Fortunak of Howard University, Washington for his expert advice and support in the estimation of the costs of API for several drugs analysed in this study.

